# Masseter Muscle Volume and Its Association with Sarcopenia and Muscle Determinants with Insights from ACTN3 Polymorphism in Older Japanese Adults: the Bunkyo Health Study

**DOI:** 10.1101/2024.04.17.24305946

**Authors:** Abulaiti Abudurezake, Saori Kakehi, Futaba Umemura, Hideyoshi Kaga, Yuki Someya, Hiroki Tabata, Yasuyo Yoshizawa, Hitoshi Naito, Tsubasa Tajima, Naoaki Ito, Hikaru Otsuka, Huicong Shi, Mari Sugimoto, Shota Sakamoto, Yukiko Muroga, Hidetaka Wakabayashi, Ryuzo Kawamori, Hirotaka Watada, Yoshifumi Tamura

## Abstract

**Aim:** Sarcopenia has been associated with a decrease in masseter muscle (MM) thickness in high-risk older populations. However, the relationship to sarcopenia and determinants of MM volume (MMV) in the general older population remain unclear.

**Mabethod:** In a cross-sectional study of 1484 older adults of Tokyo, we evaluated MMV using 3D MRI scanning, appendicular skeletal muscle mass (ASMM), handgrip strength, dietary intake, smoking, insulin-like growth factor 1 (IGF-1) levels, and the ACTN3 R577X polymorphism. Participants were divided into quintiles based on MMV (Q1-5).

**Results:** Our study of participants with a mean age of 73.0 ± 5.3 years, MMV (Men:35.3 ± 7.8 ml, Women: 25.0 ± 5.1 ml) was significantly larger in men than in women. A significant association between MMV and sarcopenia was observed, with the lowest quintile (Q1) showing a higher risk compared to the highest quintile (Q5) in both sexes. Body mass index (BMI) and age were independent determinants of ASMM in both sexes, while BMI, but interestingly not age, was a determinant of MMV. Moreover, IGF-1 was positively correlated with MMV in both sexes; smoking negatively correlated with MMV in women. The ACTN3 577X polymorphism was associated with only smaller MMV in men.

**Conclusion:** Low MMV increased the risk of sarcopenia, particularly in men. BMI and age strongly influenced ASMM, while MMV was only weakly associated with BMI and not with age. Notably, the IGF-1 level was positively correlated to only MMV, and the ACTN3 genotype was linked to reduced only MMV in men.

## Introduction

The rise in the number of older individuals requiring care is a grave issue in aging societies(1). One potential solution to this problem lies in preventative measures that reduce the risk of needing care. Sarcopenia, a condition characterized by diminished muscle mass, reduced muscle strength, and compromised physical function(2, 3), contributes significantly to the increased risk of mortality and the need for care(4, 5). Thus, it is presumed that the prevention of sarcopenia plays a pivotal role in mitigating care requirements.

Several risk factors have been associated with an increased risk of sarcopenia, including aging(6), reduced BMI(7), decreased physical activity(8), and cerebrovascular disorders(9). Two previous studies highlighted the relationship between masseter muscle (MM) thickness and sarcopenia (10, 11). In these studies, a mere 1-mm increase in MM thickness, as measured by ultrasound, significantly decreased the risk of sarcopenia by ∼45% and ∼57% in older patients hospitalized for hip fractures and those in nursing homes, respectively. In addition, MM thickness was significantly associated with skeletal mass index (SMI) in community-dwelling older people(12). Given these findings, it is reasonable to expect that decreased MM volume (MMV) may also be associated with sarcopenia in the general older population. However, the potential association between MMV and formally diagnosed sarcopenia, as defined by the Asian Working Group for Sarcopenia (AWGS) 2019 (3), in this population remains unexplored. Another issue is that the aforementioned studies measured muscle size using ultrasound, which is not the most accurate method(13). CT or MRI scans are superior because they provide detailed three-dimensional images that enable more precise measurement of muscle volume. In particular, although CT and MRI scans of the head have been performed in older people, MM has not been analyzed at all, and it would be clinically useful to identify standard MMV values and risk prediction thresholds.

Even if an association is observed between MMV and sarcopenia, the determinants of each may be different. It is rare for MM to contract in relation to physical activity or to exert maximal muscle force(14, 15), and therefore it differs from skeletal muscles of the extremities, which are active during physical activity. Therefore, comparing the determinants of appendicular skeletal muscle mass (ASMM) and MMV may provide important insights into why MMV is associated with sarcopenia, and may yield a deeper understanding of the mechanisms of muscle mass regulation.

Genetic factors are determinants of skeletal muscle mass and function(16–19), and one of the most extensively studied genes in this context is the α-actinin-3 gene. The protein α-actinin-3, coded by *ACTN3*, forms part of the Z-line of muscle fibers(20) and is specifically expressed only in fast-twitch skeletal muscle fibers(14). Notably, homozygosity for a common nonsense polymorphism (R577X) in the *ACTN3* gene results in a complete deficiency of α-actinin-3, and in animal models, the lack of actn3 leads to a reduction in both muscle mass, mainly type II fibers, and muscle strength(21). Indeed, the ACTN3 577XX α-actinin-3 null genotype is significantly underrepresented in elite sprint athletes(22), and it increases the risk of sarcopenia in older people (19, 23). However, it remains unclear whether the ACTN3 577XX null genotype is associated with reduced MMV.

Against this background, our study aimed to clarify the relationship between MMV (determined through 3D reconstruction of MRI images) and sarcopenia in older Japanese people. Furthermore, we compared the determinants of MMV and ASMM, including genetic polymorphisms of *ACTN3*.

## Research Design and Methods

### Study Design and Participants

This cross-sectional study used baseline data from the Bunkyo Health Study(1). Briefly, in this study, we recruited individuals aged between 65–84 years living in Bunkyo-ku, an urban area in Tokyo, Japan. The Bunkyo Health Study registry initially enrolled 1629 individuals. From this cohort, we excluded 64 participants who either lacked head MRI data or image slices that included the masticatory muscles. Additionally, among the remaining 1565 participants, 66 diagnosed with cerebrovascular disorders and 15 with unavailable data (body composition [n = 2], physical function [n = 3], genetic type [n = 7]) were also excluded from the study. Consequently, the final sample for this analysis comprised 1484 participants (603 men, 881 women) (Figure S1). All participants completed the examinations at the Sportology Center from October 15, 2015, to October 1, 2018. The study protocol was approved by the ethics committee of Juntendo University in November 2015 (Nos. 2015078, 2016138, 2016131, 2017121, and 2019085). This study was carried out in accordance with the principles outlined in the Declaration of Helsinki. All participants gave written informed consent and were informed that they had the right to withdraw from the trial at any time.

### 3D Assessment of MMV

Total MMV was measured by selecting and extracting the data captured by MRI with a 3D medical image analysis system, and reconstructing these data in 3D (24). MRI was performed in the supine position with the occlusion relaxed. All head MRI data (2-mm-wide T2-weighted images taken with a 0.3 T MRI, Hitachi, Ltd.) were imported into the workstation software application, AZE Medical Image Processor (AZE Virtual Place, Canon Medical Co., Ltd.). For each slice, the masticatory muscles were selected and extracted using a semi-automatic function (Figure S2). The MM was then reconstructed using the contour trace function. We performed color mapping to ensure that the entire MM was visible, and the volume was measured using the 3D volumetry function (Figure S2, 3). We used the mean value of the left and right MMVs. A.A. measured all MMVs, and the intraclass correlation coefficient and its 95% confidence interval (CI) were 0.944 and 0.989–0.997, respectively.

### Body composition measurements

ASMM was evaluated with bioelectrical impedance analysis (InBody770, InBody Japan Inc., Tokyo, Japan), and the SMI was calculated (ASMM/height^2^).

### Genotyping

Genomic DNA was isolated from whole blood using the standard method (RNeasy Blood and Tissue DNA Extraction Kit, QIAGEN, Minneapolis, MN, USA). All participants were genotyped for the ACTN3 R577X (rs1815739) polymorphism using the Infinium Asian Screening Array-24 v1.0 BeadChip (Illumina, San Diego, CA, USA).

### Other measurements

Handgrip strength (HGS) was measured using a handgrip dynamometer (T.K.K. 5401, Takei Scientific Instruments, Niigata, Japan) with the arm held to the side of the body. Physical activity level was evaluated using the International Physical Activity Questionnaire, which assesses different types of physical activity, such as walking and both moderate- and high-intensity activities(25). As part of the survey on dietary history, participants completed the brief diet history questionnaire (BDHQ)(26). Based on responses to the question about smoking, cumulative cigarette consumption was expressed as the Brinkman index (cigarette consumption per day multiplied by the number of years smoking)(27). Insulin-like growth factor 1 (IGF-1) levels were measured in blood samples collected from participants after an overnight fast.

### Definition of sarcopenia

Sarcopenia was defined as weak HGS (<28 kg for men and <18 kg for women) and low ASMM (<7.0 kg/m^2^ for men and <5.7 kg/m^2^ for women) based on the definition of the AWGS 2019 (3).

### Statistical analyses

All statistical calculations were performed using IBM SPSS Statistics for Windows version 25 (IBM, Armonk, NY). Data are presented as means ± SD or number (%). Differences between sexes were determined using Student’s t-test for continuous variables and the χ^2^ test for non-continuous variables (Table 1). Participants were divided into quintiles based on the MMV for each sex (quintiles 1-5: Q1-5). Differences in means and proportions were evaluated using ANOVA and the χ^2^ test (Table 2). Differences in characteristics between genotypes (Table 4) were compared using ANOVA and the χ^2^ test. Adjustments for multiple comparisons were performed using post hoc Bonferroni correction. Logistic regression was used to estimate odds ratios (ORs) and 95% confidence intervals (CIs) for the prevalence of each outcome in each group, compared with the reference group. Multiple linear regression was used to examine factors affecting MMV or ASMM in each group. Potential confounders were based on previous reports that have identified factors such as BMI, physical activity level, protein intake, diabetes, smoking, and IGF-1 as being associated with muscle mass or muscle strength(7, 8, 28), these factors were included as confounders in our analysis. All statistical tests were two-sided with a 5% significant level.

**Table 1.** Characteristics of the study participants.

|  | All | Men | Women | p value |
| --- | --- | --- | --- | --- |
| N | 1,484 | 603 | 881 |  |
| Age (years) | 73.0 ± 5.3 | 72.9 ± 5.3 | 73.0 ± 5.4 | 0.716 |
| Masseter muscle volume (ml) | 29.2 ± 8.1 | 35.3 ± 7.8 | 25.0 ± 5.1 | <0.001 |
| Height (cm) | 157.9 ± 8.7 | 165.8 ± 5.9 | 152.4 ± 5.5 | <0.001 |
| Weight (kg) | 56.9 ± 10.2 | 64.6 ± 8.6 | 51.7 ± 7.6 | <0.001 |
| Body mass index (kg/m<sup>2</sup>) | 22.7 ± 3.1 | 23.5 ± 2.8 | 22.2 ± 3.2 | <0.001 |
| Appendicular skeletal muscle mass (kg) | 16.2 ± 3.9 | 20.2 ± 2.6 | 13.5 ± 1.8 | <0.001 |
| Skeletal muscle index (kg/m<sup>2</sup>) | 6.4 ± 1.0 | 7.3 ± 0.6 | 5.8 ± 0.6 | <0.001 |
| Hand grip strength (kg) | 25.7 ± 7.0 | 32.4 ± 5.4 | 21.2 ± 3.5 | <0.001 |
| Total physical activity (Mets*hour/week) | 44.2 ± 48.7 | 48.3 ± 56.8 | 41.4 ± 42.0 | 0.007 |
| Low SMI (%) | 38.2 | 30.8 | 43.2 | <0.001 |
| Low hand grip strength (%) | 18.7 | 21.6 | 16.7 | 0.018 |
| Sarcopenia (%) | 12.0 | 12.4 | 11.7 | 0.664 |
| Energy intake (kcal/day) | 1959.6 ± 597.7 | 2139.7 ± 629.3 | 1836.3 ± 541.9 | <0.001 |
| Protein intake (g/day) | 83.0 ± 30.7 | 83.9 ± 30.4 | 82.3 ± 30.8 | 0.325 |
| Brinkman index | 246.8 ± 471.9 | 512.3 ± 594.0 | 65.1 ± 229.2 | <0.001 |
| IGF-1 (ng/mL) | 93.4 ± 29.4 | 98.0 ± 28.8 | 90.3 ± 29.4 | <0.001 |
| Diabetes (%) | 11.4 | 16.6 | 7.8 | <0.001 |

Data are expressed as mean ± SD. p values are derived from the Student's t-test for continuous variables and the $\chi^2$ test for non-continuous variables.

**Table 2.**
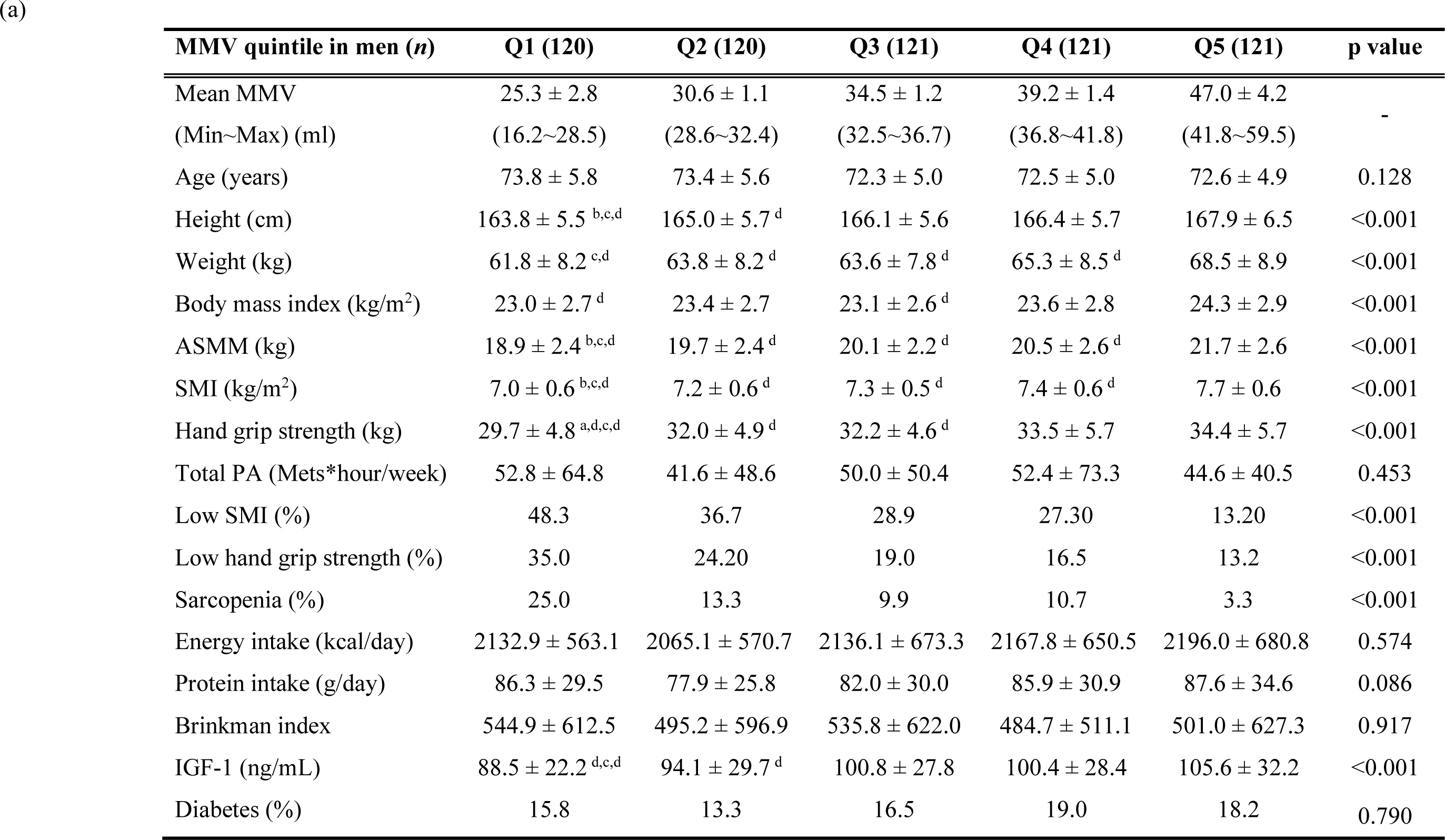
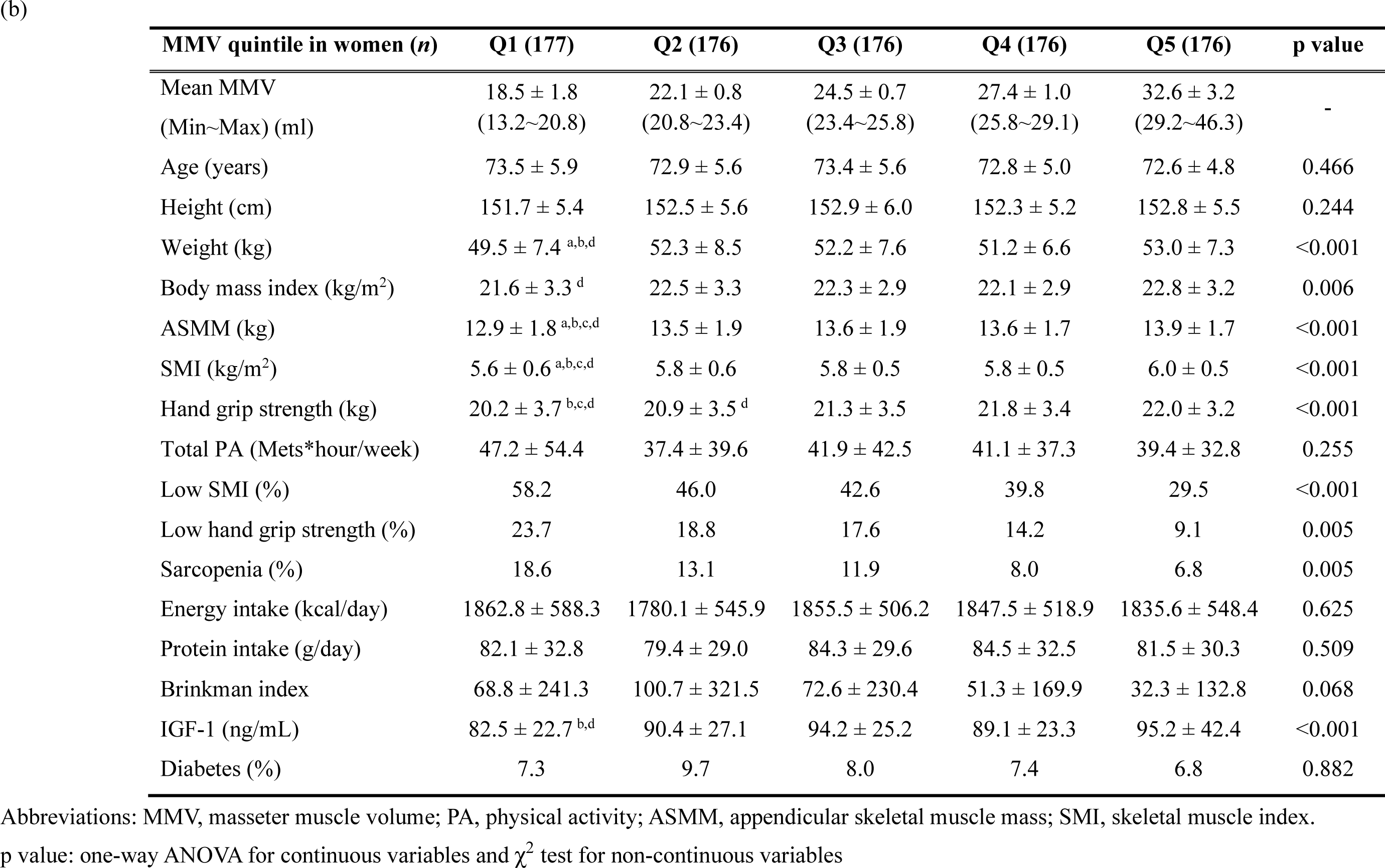
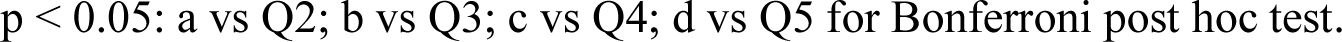
Characteristics of study participants by MMV quintiles in men (a) and women (b)

| MMV quintile in men (*n*) | Q1 (120) | Q2 (120) | Q3 (121) | Q4 (121) | Q5 (121) | p value |
| --- | --- | --- | --- | --- | --- | --- |
| Mean MMV | 25.3 ± 2.8 | 30.6 ± 1.1 | 34.5 ± 1.2 | 39.2 ± 1.4 | 47.0 ± 4.2 | - |
| (Min~Max) (ml) | (16.2~28.5) | (28.6~32.4) | (32.5~36.7) | (36.8~41.8) | (41.8~59.5) |  |
| Age (years) | 73.8 ± 5.8 | 73.4 ± 5.6 | 72.3 ± 5.0 | 72.5 ± 5.0 | 72.6 ± 4.9 | 0.128 |
| Height (cm) | 163.8 ± 5.5<sup>b,c,d</sup> | 165.0 ± 5.7<sup>d</sup> | 166.1 ± 5.6 | 166.4 ± 5.7 | 167.9 ± 6.5 | <0.001 |
| Weight (kg) | 61.8 ± 8.2<sup>c,d</sup> | 63.8 ± 8.2<sup>d</sup> | 63.6 ± 7.8<sup>d</sup> | 65.3 ± 8.5<sup>d</sup> | 68.5 ± 8.9 | <0.001 |
| Body mass index (kg/m<sup>2</sup>) | 23.0 ± 2.7<sup>d</sup> | 23.4 ± 2.7 | 23.1 ± 2.6<sup>d</sup> | 23.6 ± 2.8 | 24.3 ± 2.9 | <0.001 |
| ASMM (kg) | 18.9 ± 2.4<sup>b,c,d</sup> | 19.7 ± 2.4<sup>d</sup> | 20.1 ± 2.2<sup>d</sup> | 20.5 ± 2.6<sup>d</sup> | 21.7 ± 2.6 | <0.001 |
| SMI (kg/m<sup>2</sup>) | 7.0 ± 0.6<sup>b,c,d</sup> | 7.2 ± 0.6<sup>d</sup> | 7.3 ± 0.5<sup>d</sup> | 7.4 ± 0.6<sup>d</sup> | 7.7 ± 0.6 | <0.001 |
| Hand grip strength (kg) | 29.7 ± 4.8<sup>a,d,c,d</sup> | 32.0 ± 4.9<sup>d</sup> | 32.2 ± 4.6<sup>d</sup> | 33.5 ± 5.7 | 34.4 ± 5.7 | <0.001 |
| Total PA (Mets*hour/week) | 52.8 ± 64.8 | 41.6 ± 48.6 | 50.0 ± 50.4 | 52.4 ± 73.3 | 44.6 ± 40.5 | 0.453 |
| Low SMI (%) | 48.3 | 36.7 | 28.9 | 27.30 | 13.20 | <0.001 |
| Low hand grip strength (%) | 35.0 | 24.20 | 19.0 | 16.5 | 13.2 | <0.001 |
| Sarcopenia (%) | 25.0 | 13.3 | 9.9 | 10.7 | 3.3 | <0.001 |
| Energy intake (kcal/day) | 2132.9 ± 563.1 | 2065.1 ± 570.7 | 2136.1 ± 673.3 | 2167.8 ± 650.5 | 2196.0 ± 680.8 | 0.574 |
| Protein intake (g/day) | 86.3 ± 29.5 | 77.9 ± 25.8 | 82.0 ± 30.0 | 85.9 ± 30.9 | 87.6 ± 34.6 | 0.086 |
| Brinkman index | 544.9 ± 612.5 | 495.2 ± 596.9 | 535.8 ± 622.0 | 484.7 ± 511.1 | 501.0 ± 627.3 | 0.917 |
| IGF-1 (ng/mL) | 88.5 ± 22.2<sup>d,c,d</sup> | 94.1 ± 29.7<sup>d</sup> | 100.8 ± 27.8 | 100.4 ± 28.4 | 105.6 ± 32.2 | <0.001 |
| Diabetes (%) | 15.8 | 13.3 | 16.5 | 19.0 | 18.2 | 0.790 |

(b)
| MMV quintile in women (*n*) | Q1 (177) | Q2 (176) | Q3 (176) | Q4 (176) | Q5 (176) | p value |
| --- | --- | --- | --- | --- | --- | --- |
| Mean MMV | 18.5 ± 1.8 | 22.1 ± 0.8 | 24.5 ± 0.7 | 27.4 ± 1.0 | 32.6 ± 3.2 | - |
| (Min~Max) (ml) | (13.2~20.8) | (20.8~23.4) | (23.4~25.8) | (25.8~29.1) | (29.2~46.3) |  |
| Age (years) | 73.5 ± 5.9 | 72.9 ± 5.6 | 73.4 ± 5.6 | 72.8 ± 5.0 | 72.6 ± 4.8 | 0.466 |
| Height (cm) | 151.7 ± 5.4 | 152.5 ± 5.6 | 152.9 ± 6.0 | 152.3 ± 5.2 | 152.8 ± 5.5 | 0.244 |
| Weight (kg) | 49.5 ± 7.4<sup>a,b,d</sup> | 52.3 ± 8.5 | 52.2 ± 7.6 | 51.2 ± 6.6 | 53.0 ± 7.3 | <0.001 |
| Body mass index (kg/m<sup>2</sup>) | 21.6 ± 3.3<sup>d</sup> | 22.5 ± 3.3 | 22.3 ± 2.9 | 22.1 ± 2.9 | 22.8 ± 3.2 | 0.006 |
| ASMM (kg) | 12.9 ± 1.8<sup>a,b,c,d</sup> | 13.5 ± 1.9 | 13.6 ± 1.9 | 13.6 ± 1.7 | 13.9 ± 1.7 | <0.001 |
| SMI (kg/m<sup>2</sup>) | 5.6 ± 0.6<sup>a,b,c,d</sup> | 5.8 ± 0.6 | 5.8 ± 0.5 | 5.8 ± 0.5 | 6.0 ± 0.5 | <0.001 |
| Hand grip strength (kg) | 20.2 ± 3.7<sup>b,c,d</sup> | 20.9 ± 3.5<sup>d</sup> | 21.3 ± 3.5 | 21.8 ± 3.4 | 22.0 ± 3.2 | <0.001 |
| Total PA (Mets*hour/week) | 47.2 ± 54.4 | 37.4 ± 39.6 | 41.9 ± 42.5 | 41.1 ± 37.3 | 39.4 ± 32.8 | 0.255 |
| Low SMI (%) | 58.2 | 46.0 | 42.6 | 39.8 | 29.5 | <0.001 |
| Low hand grip strength (%) | 23.7 | 18.8 | 17.6 | 14.2 | 9.1 | 0.005 |
| Sarcopenia (%) | 18.6 | 13.1 | 11.9 | 8.0 | 6.8 | 0.005 |
| Energy intake (kcal/day) | 1862.8 ± 588.3 | 1780.1 ± 545.9 | 1855.5 ± 506.2 | 1847.5 ± 518.9 | 1835.6 ± 548.4 | 0.625 |
| Protein intake (g/day) | 82.1 ± 32.8 | 79.4 ± 29.0 | 84.3 ± 29.6 | 84.5 ± 32.5 | 81.5 ± 30.3 | 0.509 |
| Brinkman index | 68.8 ± 241.3 | 100.7 ± 321.5 | 72.6 ± 230.4 | 51.3 ± 169.9 | 32.3 ± 132.8 | 0.068 |
| IGF-1 (ng/mL) | 82.5 ± 22.7<sup>b,d</sup> | 90.4 ± 27.1 | 94.2 ± 25.2 | 89.1 ± 23.3 | 95.2 ± 42.4 | <0.001 |
| Diabetes (%) | 7.3 | 9.7 | 8.0 | 7.4 | 6.8 | 0.882 |

Abbreviations: MMV, masseter muscle volume; PA, physical activity; ASMM, appendicular skeletal muscle mass; SMI, skeletal muscle index.
p value: one-way ANOVA for continuous variables and $\chi^2$ test for non-continuous variables
$p < 0.05$ : a vs Q2; b vs Q3; c vs Q4; d vs Q5 for Bonferroni post hoc test.

**Table 3.**
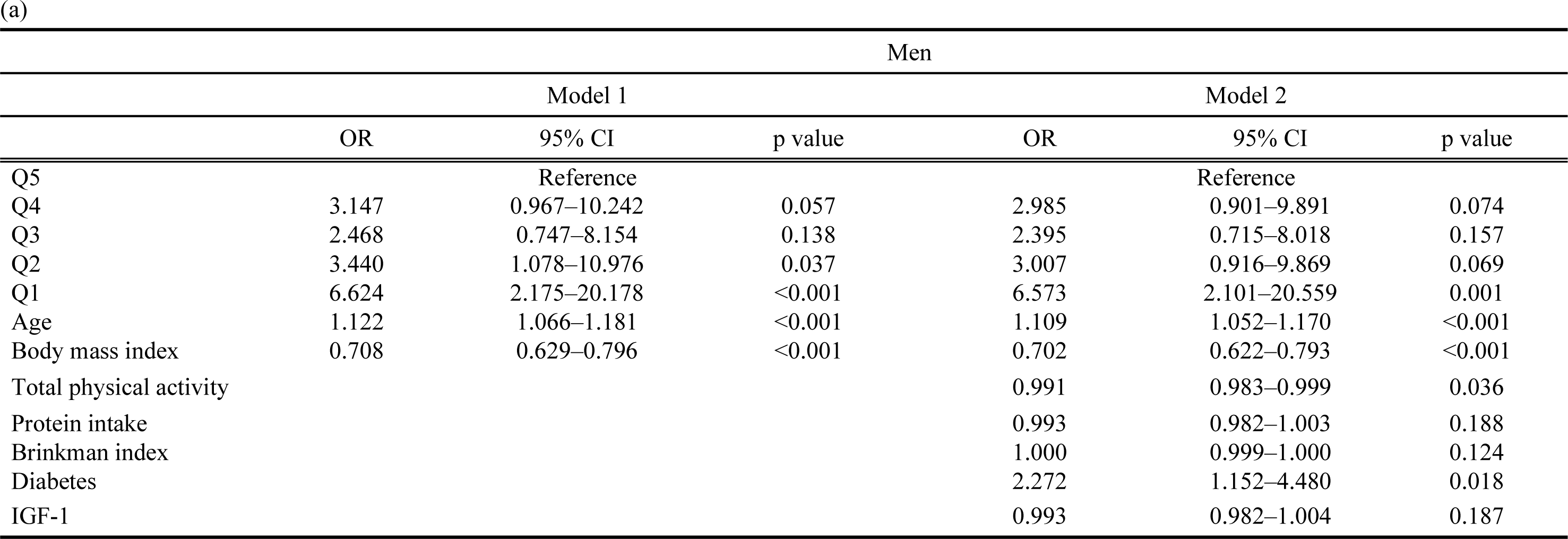
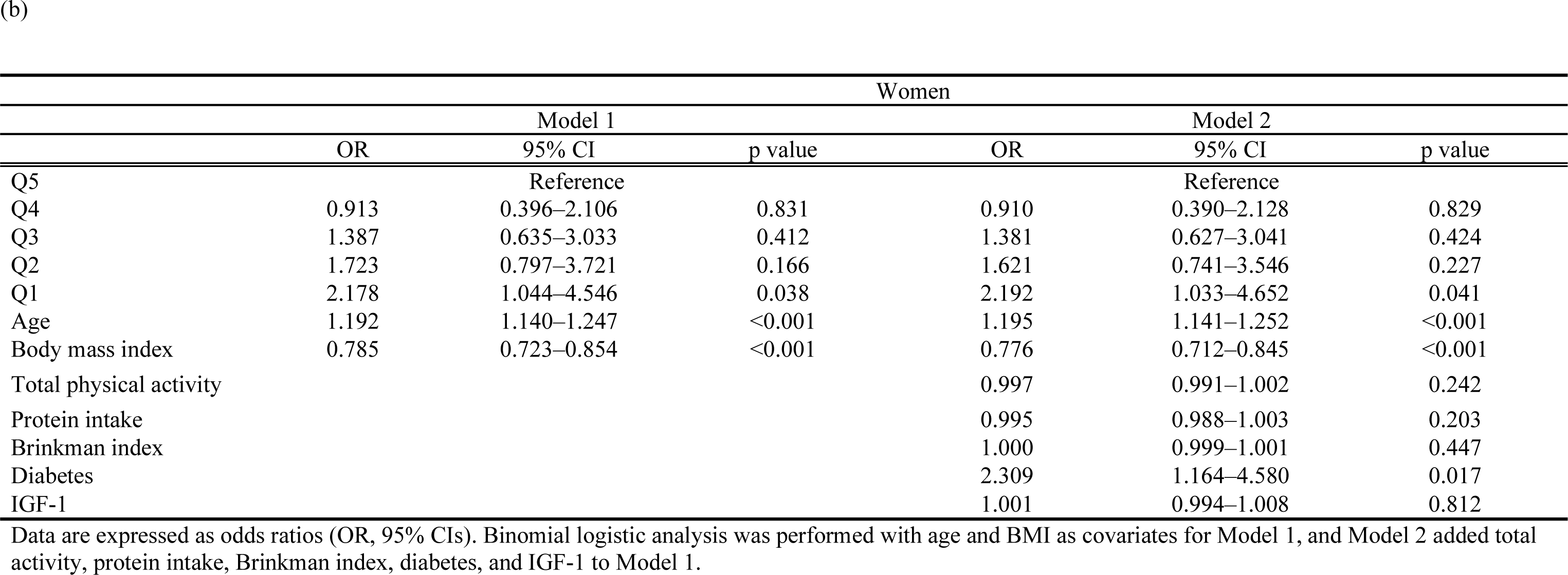
Odds ratios for sarcopenia by MMV quintiles in men (a) and women (b): results of a binomial logistic regression analysis. (a)

**Table 4.**
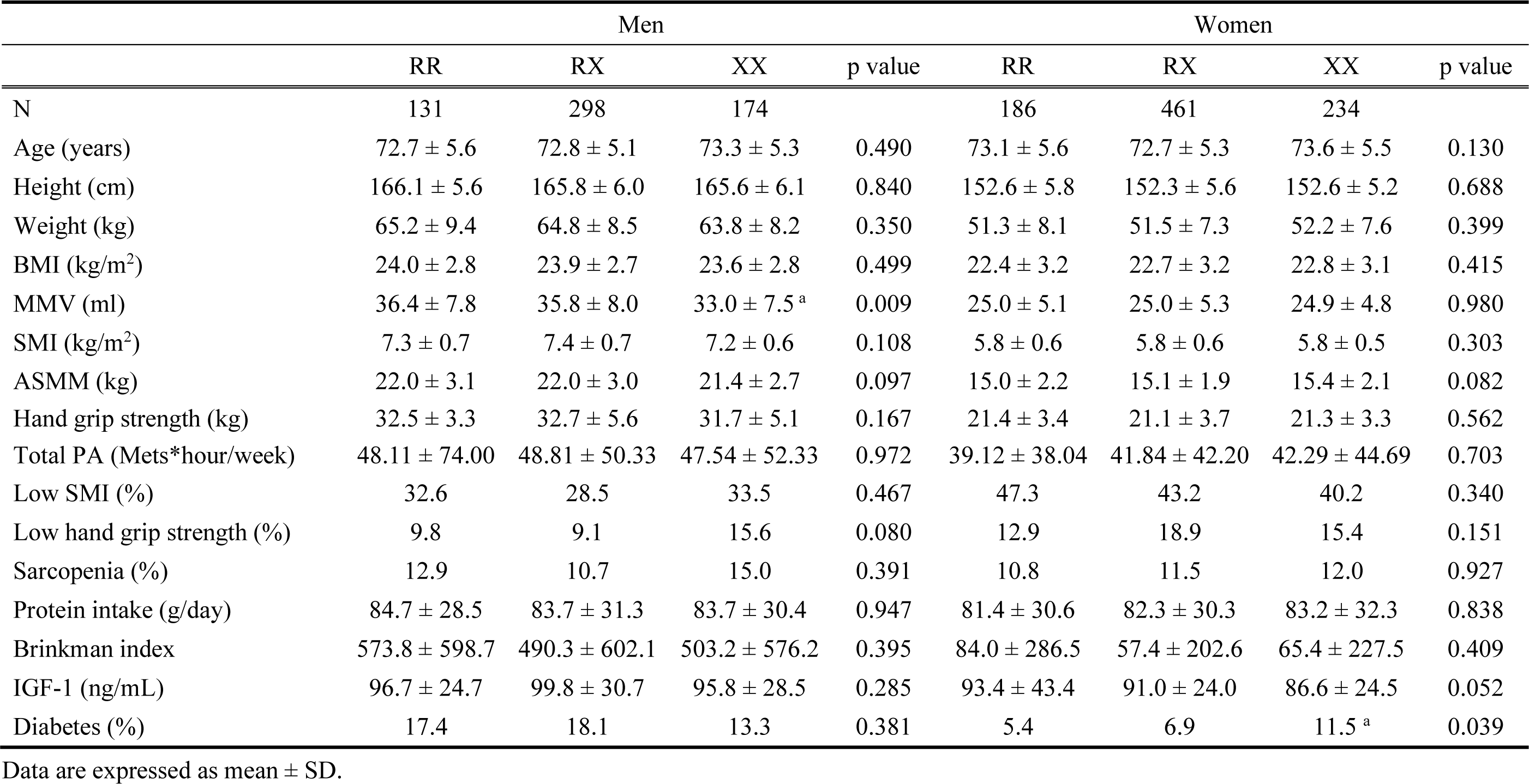
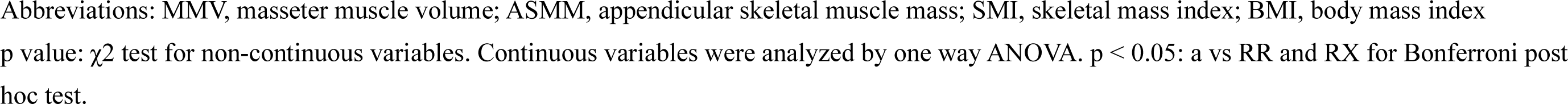
Characteristics of older participants by ACTN3 R577X genotype.

|  | Men |  |  |  | Women |  |  |  |
| --- | --- | --- | --- | --- | --- | --- | --- | --- |
|  | RR | RX | XX | p value | RR | RX | XX | p value |
| N | 131 | 298 | 174 |  | 186 | 461 | 234 |  |
| Age (years) | 72.7 ± 5.6 | 72.8 ± 5.1 | 73.3 ± 5.3 | 0.490 | 73.1 ± 5.6 | 72.7 ± 5.3 | 73.6 ± 5.5 | 0.130 |
| Height (cm) | 166.1 ± 5.6 | 165.8 ± 6.0 | 165.6 ± 6.1 | 0.840 | 152.6 ± 5.8 | 152.3 ± 5.6 | 152.6 ± 5.2 | 0.688 |
| Weight (kg) | 65.2 ± 9.4 | 64.8 ± 8.5 | 63.8 ± 8.2 | 0.350 | 51.3 ± 8.1 | 51.5 ± 7.3 | 52.2 ± 7.6 | 0.399 |
| BMI (kg/m<sup>2</sup>) | 24.0 ± 2.8 | 23.9 ± 2.7 | 23.6 ± 2.8 | 0.499 | 22.4 ± 3.2 | 22.7 ± 3.2 | 22.8 ± 3.1 | 0.415 |
| MMV (ml) | 36.4 ± 7.8 | 35.8 ± 8.0 | 33.0 ± 7.5 <sup>a</sup> | 0.009 | 25.0 ± 5.1 | 25.0 ± 5.3 | 24.9 ± 4.8 | 0.980 |
| SMI (kg/m<sup>2</sup>) | 7.3 ± 0.7 | 7.4 ± 0.7 | 7.2 ± 0.6 | 0.108 | 5.8 ± 0.6 | 5.8 ± 0.6 | 5.8 ± 0.5 | 0.303 |
| ASMM (kg) | 22.0 ± 3.1 | 22.0 ± 3.0 | 21.4 ± 2.7 | 0.097 | 15.0 ± 2.2 | 15.1 ± 1.9 | 15.4 ± 2.1 | 0.082 |
| Hand grip strength (kg) | 32.5 ± 3.3 | 32.7 ± 5.6 | 31.7 ± 5.1 | 0.167 | 21.4 ± 3.4 | 21.1 ± 3.7 | 21.3 ± 3.3 | 0.562 |
| Total PA (Mets*hour/week) | 48.11 ± 74.00 | 48.81 ± 50.33 | 47.54 ± 52.33 | 0.972 | 39.12 ± 38.04 | 41.84 ± 42.20 | 42.29 ± 44.69 | 0.703 |
| Low SMI (%) | 32.6 | 28.5 | 33.5 | 0.467 | 47.3 | 43.2 | 40.2 | 0.340 |
| Low hand grip strength (%) | 9.8 | 9.1 | 15.6 | 0.080 | 12.9 | 18.9 | 15.4 | 0.151 |
| Sarcopenia (%) | 12.9 | 10.7 | 15.0 | 0.391 | 10.8 | 11.5 | 12.0 | 0.927 |
| Protein intake (g/day) | 84.7 ± 28.5 | 83.7 ± 31.3 | 83.7 ± 30.4 | 0.947 | 81.4 ± 30.6 | 82.3 ± 30.3 | 83.2 ± 32.3 | 0.838 |
| Brinkman index | 573.8 ± 598.7 | 490.3 ± 602.1 | 503.2 ± 576.2 | 0.395 | 84.0 ± 286.5 | 57.4 ± 202.6 | 65.4 ± 227.5 | 0.409 |
| IGF-1 (ng/mL) | 96.7 ± 24.7 | 99.8 ± 30.7 | 95.8 ± 28.5 | 0.285 | 93.4 ± 43.4 | 91.0 ± 24.0 | 86.6 ± 24.5 | 0.052 |
| Diabetes (%) | 17.4 | 18.1 | 13.3 | 0.381 | 5.4 | 6.9 | 11.5 <sup>a</sup> | 0.039 |

Data are expressed as mean ± SD.
Abbreviations: MMV, masseter muscle volume; ASMM, appendicular skeletal muscle mass; SMI, skeletal mass index; BMI, body mass index p value: $\chi^2$ test for non-continuous variables. Continuous variables were analyzed by one way ANOVA. $p < 0.05$ : a vs RR and RX for Bonferroni post hoc test.

## Results

### Characteristics of study participants

Characteristics of the study participants are shown in Table 1. Of 1,484 participants (average age 73.0 ± 5.3 years), 603 were men (average age 72.9 ± 5.3 years) and 881 were women (average age 73.0 ± 5.4 years). The MMV and SMI were about 1.4 and 1.3 times greater in men than in women, respectively, indicating a significant difference in both cases. HGS was 1.5 times greater in men than in women, also a significant difference. The prevalence of diabetes was significantly higher in men at 16.5% compared to women at 7.8%. The prevalence of sarcopenia was comparable between men and women, at 12.4% in men and 11.7% in women.

### Association between MMV and various body composition indices

MMV was divided into quintiles and clinical features were compared (Table 2). In men, Q1, representing the lowest MMV, was associated with the lowest values among the groups for all clinical features other than age and physical activity, and was associated with the highest proportions of participants with low SMI, low HGS, and sarcopenia. Compared to Q1, there were significant differences between groups in terms of height (against Q3, 4, 5), weight (against Q4, 5), BMI (against Q5), ASMM (against Q3, 4, 5), SMI (against Q3, 4, 5), HGS (against Q2, 3, 4, 5), and IGF-1 (for Q3, 4, 5). Similar trends were observed in women. Compared to Q1, there were significant differences between groups in weight (against Q2, 3, 5), BMI (against Q5), ASMM (against Q2, 3, 4, 5), SMI (against Q2, 3, 4, 5), HGS (against Q3, 4, 5), and IGF-1 (for Q3, 5).

### Relationship between MMV and sarcopenia

To clarify whether decreased MMV was independently associated with the prevalence of sarcopenia, we conducted a logistic regression analysis (Table 3). After adjusting for age and BMI in Model 1, Q1 was associated with a significantly higher OR for the prevalence of sarcopenia compared to Q5 in both men and women (men, OR: 6.624, 95% CI: 2.175– 20.178, p < 0.001; women, OR: 2.178, 95% CI: 1.044–4.546, p = 0.038), and the OR for Q2 in men was also significantly different relative to Q5 (OR: 3.440, 95% CI: 1.078– 10.976, p = 0.037). In model 2, which adjusted for physical activity, protein intake, Brinkman index, and IGF-1, the ORs of Q1 in men (OR: 6.573, 95% CI: 2.101–20.559, p = 0.001) and in women (OR: 2.192, 95% CI: 1.033–4.652, p = 0.041) were still significant compared to Q5.

### Relationship between ACTN3 R577X polymorphism and MMV

Next, we examined the association between MMV or ASMM and ACTN3 577XX genotypes. The samples exhibited no deviation from the Hardy-Weinberg equilibrium. As shown in Table 4, in men with the ACTN3 577XX genotype, MMV was significantly lower compared to other genotypes, but ASMM was similar across genotypes. In contrast, no association was found between genotype and either MMV or ASMM in women.

### Multiple regression model of factors predicting MMV or ASMM

We next performed a multiple regression analysis to identify the independent determinants of MMV and ASMM (Table 5). In the total variance explained by the model, *R*^2^ was much lower in MMV than in ASMM in both sexes. This difference was mainly due to the much more significant standardized β values of BMI and age for ASMM than for MMV in both sexes. BMI was positively correlated (men, β = 0.618; women, β = 0.600) and age was negatively correlated (men, β = -0.296; women, β = 0.246) with ASMM in both sexes, while BMI (men, β = -0.168; women, β = -0.110), but interestingly not age, was positively correlated with MMV. In addition, IGF-1 was positively correlated with MMV in both sexes, while smoking was negatively correlated with MMV in women. The ACTN3 577X polymorphism in men, but not in women, was associated with smaller MMV, which was consistent with the analysis in Table 4.

**Table 5.** Association of each factor with MMV or ASMM in a multiple regression model.

|  | Men |  |  |  | Women |  |  |  |
| --- | --- | --- | --- | --- | --- | --- | --- | --- |
|  | MMV |  | ASMM |  | MMV |  | ASMM |  |
| | $\beta$ | p value | $\beta$ | p value | $\beta$ | p value | $\beta$ | p value |
| actn3 | -0.100 | 0.011 | -0.032 | 0.280 | 0.003 | 0.940 | 0.058 | 0.058 |
| Age | -0.047 | 0.250 | -0.296 | <0.001 | -0.048 | 0.173 | -0.246 | <0.001 |
| Body mass index | 0.168 | <0.001 | 0.618 | <0.001 | 0.110 | 0.001 | 0.600 | <0.001 |
| Total physical activity | -0.016 | 0.685 | 0.017 | 0.578 | -0.020 | 0.549 | -0.004 | 0.870 |
| Protein intake | 0.044 | 0.272 | 0.017 | 0.581 | 0.042 | 0.217 | 0.048 | 0.076 |
| Brinkman index | -0.023 | 0.573 | 0.002 | 0.949 | -0.072 | 0.032 | -0.019 | 0.488 |
| Diabetes | 0.060 | 0.134 | 0.011 | 0.721 | -0.011 | 0.745 | -0.031 | 0.250 |
| IGF-1 | 0.173 | <0.001 | 0.028 | 0.366 | 0.129 | <0.001 | 0.042 | 0.128 |
| $R^2$ | 0.086* | | 0.474* | | 0.042* | | 0.391* | |

\*p < 0.001 for analysis of variance

## Discussion

In the present study, we measured MMV in community-dwelling older people in Japan and investigated its relationship with sarcopenia. Furthermore, we compared determinants of MMV and ASMM, including genetic polymorphisms of *ACTN3*. Our findings indicated that men in the group with the lowest MMV had a 6.9-fold increased risk of sarcopenia, while women had a 2.1-fold increased risk, compared to those in the group with the highest MMV. Multiple regression analysis revealed that both BMI and age were independent determinants of ASMM in both sexes. However, for MMV, the *ACTN3* genotype and IGF-1 were independent determinants in men, whereas smoking and IGF-1 emerged as significant factors in women.

The present study revealed an association between MMV and sarcopenia, which is consistent with the findings of prior cohort studies(10, 11) that utilized ultrasound to measure MMT. Importantly, our participants were community-dwelling individuals who were over a decade younger (73 years old) compared to those in previous studies (86 or 84.7 years old), and were either hospitalized for hip fractures or who resided in nursing homes(10, 11). In this large-scale study, we found a substantial rise in the OR of sarcopenia in men within the two lowest quintiles of MMV (Q1 and Q2) when compared to the group with the largest MMV (Q5). Conversely, in women, a significant increase in OR was only observed in Q1, although it was not as pronounced as in men. Moreover, the OR gradually increased as MMV decreased in women, while in men, a mild although not significant rise was detected in Q3 and Q4. Thus, despite gender differences, a trend toward an increased risk of sarcopenia with decreasing MMV was observed in both men and women.

This study revealed specific differences were observed between men and women regarding the relationship between MMV and sarcopenia. In fact, after full adjustment, the OR for the lowest quintile (Q1) in men (OR: 6.573) was 3 times higher than that in women (OR: 2.192). This might be because men and women have different types of muscle fibers in their MMs. Age-related muscle atrophy occurs preferentially in type II muscle fibers(29), and it is known that the proportion of type II muscle fibers in the MM is higher in men than in women(30). Additionally, age-related loss of muscle mass is also more likely to occur in men(31). Given these factors, it is plausible that MMV serves as a more sensitive indicator of age-related muscle atrophy in men compared to women.

Previous studies have investigated the link between ACTN3 R577X and muscle mass in older people, but the results have been inconsistent. These discrepancies might arise from variations in sample sizes or the diverse ages and backgrounds of the participants. While some studies(19, 32, 33) found that people with the ACTN3 577XX null genotype were more likely to lose muscle mass, others did not observe this link(34–36). The present study did not find that ACTN3 R577X was related to ASMM, but it was notably associated with MMV, particularly in men. The α-actinin-3 protein, encoded by *ACTN3*, is specifically expressed in fast-twitch skeletal muscle fibers(14), and the proportion of type II muscle fibers in MM is higher in men than in women(30). In addition, the lack of Actn3 leads to a reduction in muscle mass in mice, predominantly in type II fibers(21), and the muscle mass reduction with age was also enhanced in Actn3-deficient mice, especially males(37). These findings may explain why there was an association between MMV and ACTN3R577X in men but not women. The lack of a clear association between ACTN3 R577X and ASMM, in contrast to MMV, may arise because factors such as BMI strongly influence ASMM, masking potential genetic differences. Similarly, even though IGF-1 is known to induce skeletal muscle hypertrophy(38), in our study it was associated with MMV, not ASMM, possibly due to the overriding effect of BMI on ASMM.

This study identified BMI and age as major determinants of ASMM. Interestingly, however, while the association between BMI and MMV was moderate, that between age and MMV was not significant. This discrepancy can be attributed to the functional differences between the two muscle types. ASM contains a higher proportion of antigravity muscles and is influenced by weight-bearing, whereas MM is primarily active during mastication. The increased weight-bearing due to a higher BMI can lead to an increase in ASMM. It is known that high weight-bearing loads predominantly result in hypertrophy of type II muscle fibers, which are more susceptible to age-related atrophy(29). In contrast, studies have shown that MM contains a higher percentage of type I fibers compared to ASM(39). We hypothesize that these differences in fiber type composition and function might explain why age affects ASM, but not MMV.

This study highlights the importance of MMV as a key marker for sarcopenia risk, particularly among older Japanese men. Since CT and MRI scans of the head are occasionally performed in the older people, it would be clinically useful to identify sarcopenia risk by performing an additional MMV analysis. In addition, clinicians should note the differential impact of BMI and age on ASMM compared to MMV, with the latter showing no association with age. Additionally, the ACTN3 577X genotype presents a unique genetic dimension, influencing MMV predominantly in men. Understanding these facts can facilitate more precise early detection of sarcopenia, as well as interventions tailored to specific gender and genetic profiles.

This study has several limitations. Because this cohort included only people living in urban areas of Japan, our results may not be applicable to people living in other parts of Japan or in other countries. In addition, since this was a cross-sectional study, we could not establish any causal relationships. Therefore, further prospective longitudinal studies are needed to clarify the association between MMV and the development of sarcopenia. In addition, since we were unable to obtain information on the remaining teeth, denture status, existence of temporomandibular joint disorder, and bruxism existence of the participants in this study. Accordingly, recent data suggested that the ACTN3 R557X polymorphism may influence bruxism in patients with dentofacial deformity(40). Therefore, further studies addressing these factors are needed.

In conclusion, low MMV was a risk factor for sarcopenia, with the effect being more pronounced in men than in women. While BMI and age were strong determinants of ASMM, only a weak association was observed between BMI and MMV, with no correlation between age and MMV. Notably, the IGF-1 level was positively correlated to MMV, but not ASMM, and the ACTN3 577X genotype linked to reduced MMV in men but showed no association with ASMM in either sex.

## Data Availability

All data produced in the present study are available upon reasonable request to the authors

## Acknowledgments

The authors would like to thank all staff for their contributions to data collection at the Sportology Center.

## Supplemental Figure legends

Figure S1 Flow chart of subject inclusion and exclusion criteria.

Figure S2 Steps for the assessment of MMV. All head MRI data are imported into the workstation software. The masseter muscle is selected and extracted. (a) The color mapping operation screen is shown. (b) The diaphragm is reconstructed. (c) The volume is measured.

Figure S3 Masseter muscle reconstructed after selective extraction.

## (References)

1. Someya Y, Tamura Y, Kaga H, Nojiri S, Shimada K, Daida H, et al. Skeletal muscle function and need for long-term care of urban elderly people in Japan (the Bunkyo Health Study): a prospective cohort study. BMJ Open. 2019;9(9):e031584.

2. Cruz-Jentoft AJ, Baeyens JP, Bauer JM, Boirie Y, Cederholm T, Landi F, et al. Sarcopenia: European consensus on definition and diagnosis: Report of the European Working Group on Sarcopenia in Older People. Age and Ageing. 2010;39(4):412–23.

3. Chen LK, Woo J, Assantachai P, Auyeung TW, Chou MY, Iijima K, et al. Asian Working Group for Sarcopenia: 2019 Consensus Update on Sarcopenia Diagnosis and Treatment. J Am Med Dir Assoc. 2020;21(3):300–7.e2.

4. Sawaya Y, Ishizaka M, Kubo A, Shiba T, Hirose T, Onoda K, et al. The Asian working group for sarcopenia’s new criteria updated in 2019 causing a change in sarcopenia prevalence in Japanese older adults requiring long-term care/support. Journal of Physical Therapy Science. 2020;32(11):742–7.

5. Ping Liu QH, Shan Hai, Hui Wang, Li Cao and B Dong. Sarcopenia as a predictor of all-cause mortality among community-dwelling older people: A systematic review and meta-analysis - ClinicalKey. Maturitas. 2017;103:16–22.

6. Wu LC, Kao HH, Chen HJ, Huang PF. Preliminary screening for sarcopenia and related risk factors among the elderly. Medicine (Baltimore). 2021;100(19):e25946.

7. Nakanishi S, Iwamoto M, Shinohara H, Iwamoto H, Kaneto H. Significance of body mass index for diagnosing sarcopenia is equivalent to slow gait speed in Japanese individuals with type 2 diabetes: Cross-sectional study using outpatient clinical data. J Diabetes Investig. 2021;12(3):417–24.

8. Kitamura M, Izawa KP, Ishihara K, Matsuda H, Okamura S, Fujioka K. Physical Activity and Sarcopenia in Community-Dwelling Older Adults with Long-Term Care Insurance. European Journal of Investigation in Health, Psychology and Education. 2021;11(4):1610–8.

9. Li W, Yue T, Liu Y. New understanding of the pathogenesis and treatment of stroke-related sarcopenia. Biomed Pharmacother. 2020;131:110721.

10. González-Fernández M, Perez-Nogueras J, Serrano-Oliver A, Torres-Anoro E, Sanz-Arque A, Arbones-Mainar JM, et al. Masseter Muscle Thickness Measured by Ultrasound as a Possible Link with Sarcopenia, Malnutrition and Dependence in Nursing Homes. Diagnostics. 2021;11(9):1587.

11. Sanz-Paris A, González-Fernandez M, Hueso-Del Río LE, Ferrer-Lahuerta E, Monge-Vazquez A, Losfablos-Callau F, et al. Muscle Thickness and Echogenicity Measured by Ultrasound Could Detect Local Sarcopenia and Malnutrition in Older Patients Hospitalized for Hip Fracture. Nutrients. 2021;13(7):2401.

12. Umeki K, Watanabe Y, Hirano H, Edahiro A, Ohara Y, Yoshida H, et al. The relationship between masseter muscle thickness and appendicular skeletal muscle mass in Japanese community-dwelling elders: A cross-sectional study. Arch Gerontol Geriatr. 2018;78:18–22.

13. Reis Durão AP, Morosolli A, Brown J, Jacobs R. Masseter muscle measurement performed by ultrasound: a systematic review. Dentomaxillofacial Radiology. 2017;46(6):20170052.

14. Miyamoto K, Ishizuka Y, Tanne K. Changes in masseter muscle activity during orthodontic treatment evaluated by a 24-hour EMG system. Angle Orthod. 1996;66(3):223–8.

15. Mew JR. The postural basis of malocclusion: a philosophical overview. Am J Orthod Dentofacial Orthop. 2004;126(6):729–38.

16. Huang Y, Bodnar D, Chen CY, Sanchez-Andrade G, Sanderson M, Biogen Biobank T, et al. Rare genetic variants impact muscle strength. Nat Commun. 2023;14(1):3449.

17. Roth SM. Genetic aspects of skeletal muscle strength and mass with relevance to sarcopenia. Bonekey Rep. 2012;1:58.

18. Charlier R, Caspers M, Knaeps S, Mertens E, Lambrechts D, Lefevre J, et al. Limited potential of genetic predisposition scores to predict muscle mass and strength performance in Flemish Caucasians between 19 and 73 years of age. Physiol Genomics. 2017;49(3):160–6.

19. Cho J, Lee I, Kang H. ACTN3 Gene and Susceptibility to Sarcopenia and Osteoporotic Status in Older Korean Adults. Biomed Res Int. 2017;2017:4239648.

20. Beggs AH, Byers TJ, Knoll JH, Boyce FM, Bruns GA, Kunkel LM. Cloning and characterization of two human skeletal muscle alpha-actinin genes located on chromosomes 1 and 11. J Biol Chem. 1992;267(13):9281–8.

21. MacArthur DG, Seto JT, Chan S, Quinlan KG, Raftery JM, Turner N, et al. An Actn3 knockout mouse provides mechanistic insights into the association between alpha-actinin-3 deficiency and human athletic performance. Hum Mol Genet. 2008;17(8):1076–86.

22. North KN, Yang N, Wattanasirichaigoon D, Mills M, Easteal S, Beggs AH. A common nonsense mutation results in alpha-actinin-3 deficiency in the general population. Nat Genet. 1999;21(4):353–4.

23. Kiuchi Y, Makizako H, Nakai Y, Taniguchi Y, Tomioka K, Sato N, et al. Associations of alpha-actinin-3 genotype with thigh muscle volume and physical performance in older adults with sarcopenia or pre-sarcopenia. Exp Gerontol. 2021;154:111525.

24. Abudurezake A, Morita T, Mori T, Amano A. Validity of Diaphragm Volume Measurements Using Three-Dimensional Computed Tomography. Juntendo Medical Journal. 2022;68(5):481–90.

25. Craig CL, Marshall AL, Sjostrom M, Bauman AE, Booth ML, Ainsworth BE, et al. International physical activity questionnaire: 12-country reliability and validity. Med Sci Sports Exerc. 2003;35(8):1381–95.

26. Kobayashi S, Murakami K, Sasaki S, Okubo H, Hirota N, Notsu A, et al. Comparison of relative validity of food group intakes estimated by comprehensive and brief-type self-administered diet history questionnaires against 16 d dietary records in Japanese adults. Public Health Nutr. 2011;14(7):1200–11.

27. Brinkman GL, Coates EO, Jr. The effect of bronchitis, smoking, and occupation on ventilation. Am Rev Respir Dis. 1963;87:684–93.

28. Roh E, Choi KM. Health Consequences of Sarcopenic Obesity: A Narrative Review. Front Endocrinol (Lausanne). 2020;11:332.

29. Ciciliot S, Rossi AC, Dyar KA, Blaauw B, Schiaffino S. Muscle type and fiber type specificity in muscle wasting. Int J Biochem Cell Biol. 2013;45(10):2191–9.

30. Tuxen A, Bakke M, Pinholt EM. Comparative data from young men and women on masseter muscle fibres, function and facial morphology. Archives of Oral Biology. 1999;44(6):509–17.

31. Goodpaster BH, Park SW, Harris TB, Kritchevsky SB, Nevitt M, Schwartz AV, et al. The loss of skeletal muscle strength, mass, and quality in older adults: the health, aging and body composition study. J Gerontol A Biol Sci Med Sci. 2006;61(10):1059–64.

32. Zempo H, Tanabe K, Murakami H, Iemitsu M, Maeda S, Kuno S. ACTN3 polymorphism affects thigh muscle area. Int J Sports Med. 2010;31(2):138–42.

33. Kahraman M, Ozulu Turkmen B, Bahat-Ozturk G, Catikkas NM, Oren MM, Sahin A, et al. Is there a relationship between ACTN3 R577X gene polymorphism and sarcopenia? Aging Clin Exp Res. 2022;34(4):757–65.

34. Bustamante-Ara N, Santiago C, Verde Z, Yvert T, Gomez-Gallego F, Rodriguez-Romo G, et al. ACE and ACTN3 genes and muscle phenotypes in nonagenarians. Int J Sports Med. 2010;31(4):221–4.

35. San Juan AF, Gomez-Gallego F, Canete S, Santiago C, Perez M, Lucia A. Does complete deficiency of muscle alpha actinin 3 alter functional capacity in elderly women? A preliminary report. Br J Sports Med. 2006;40(1):e1.

36. MacArthur DG, North KN. ACTN3: A genetic influence on muscle function and athletic performance. Exerc Sport Sci Rev. 2007;35(1):30–4.

37. Seto JT, Chan S, Turner N, MacArthur DG, Raftery JM, Berman YD, et al. The effect of alpha-actinin-3 deficiency on muscle aging. Exp Gerontol. 2011;46(4):292–302.

38. Sartori R, Romanello V, Sandri M. Mechanisms of muscle atrophy and hypertrophy: implications in health and disease. Nat Commun. 2021;12(1):330.

39. Sciote JJ, Rowlerson AM, Hopper C, Hunt NP. Fibre type classification and myosin isoforms in the human masseter muscle. J Neurol Sci. 1994;126(1):15–24.

40. Nicot R, Raoul G, Vieira AR, Ferri J, Sciote JJ. ACTN3 genotype influences masseter muscle characteristics and self-reported bruxism. Oral Dis. 2023;29(1):232–44.

