## Supplemental Figure1-3 for "Masseter Muscle Volume and Its Association with Sarcopenia and Muscle Determinants with Insights from ACTN3 Polymorphism in Older Japanese Adults: the Bunkyo Health Study"

### Slide 1
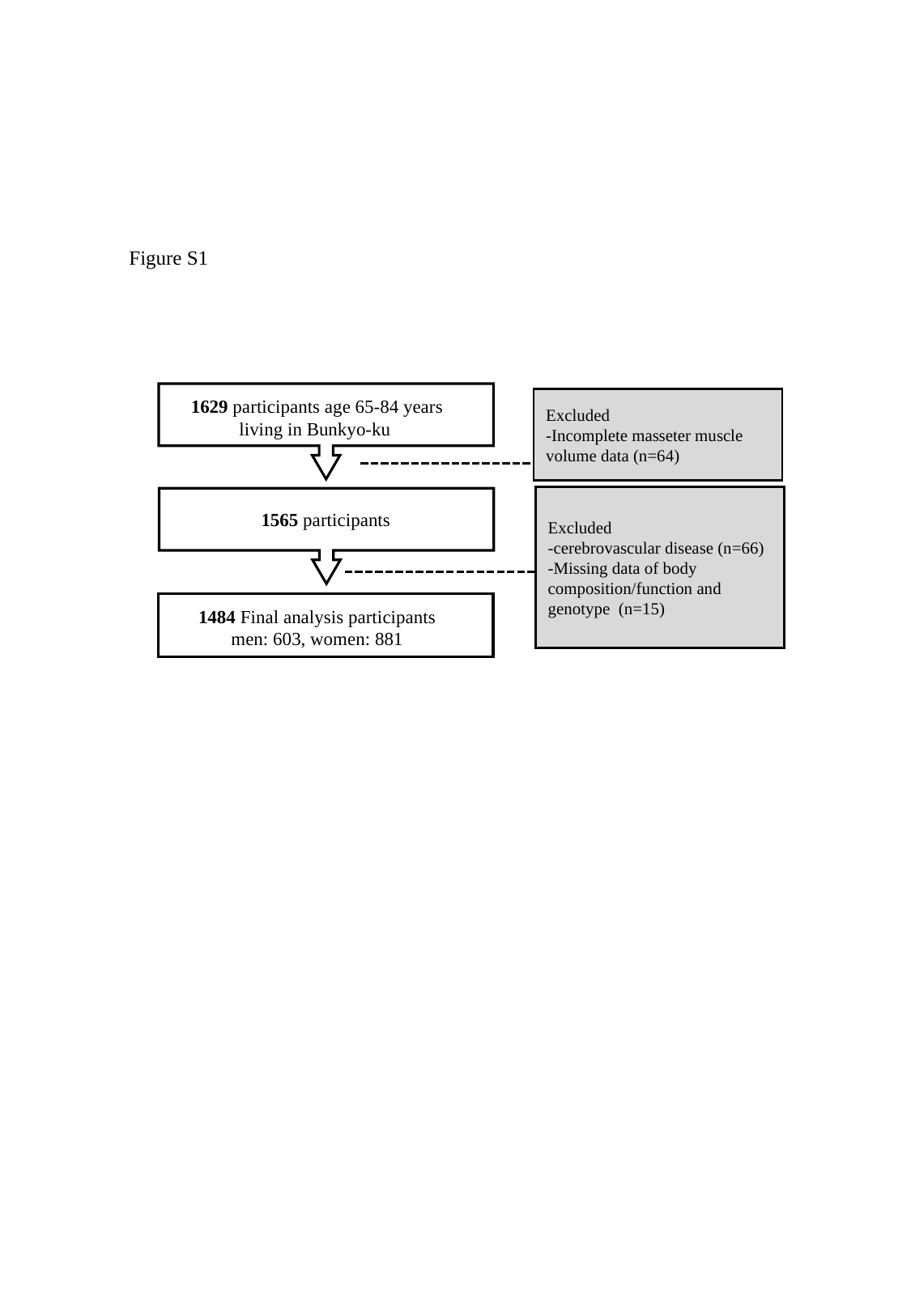

Figure S1
1629 participants age 65-84 years living in Bunkyo-ku
Excluded
-Incomplete masseter muscle volume data (n=64)
Excluded
-cerebrovascular disease (n=66)
-Missing data of body composition/function and genotype (n=15)
1565 participants
1484 Final analysis participants
men: 603, women: 881

### Slide 2
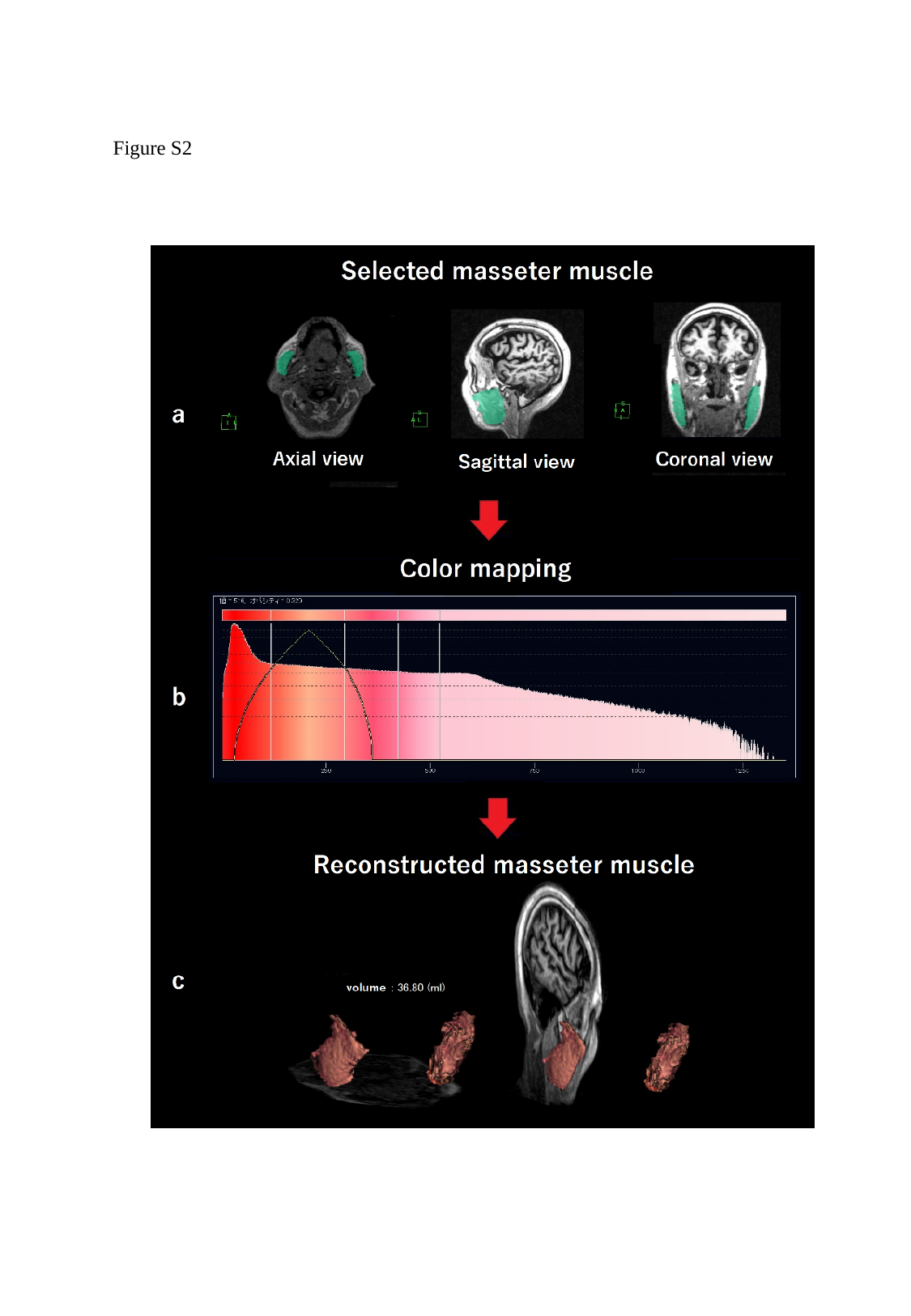

Figure S2

### Slide 3
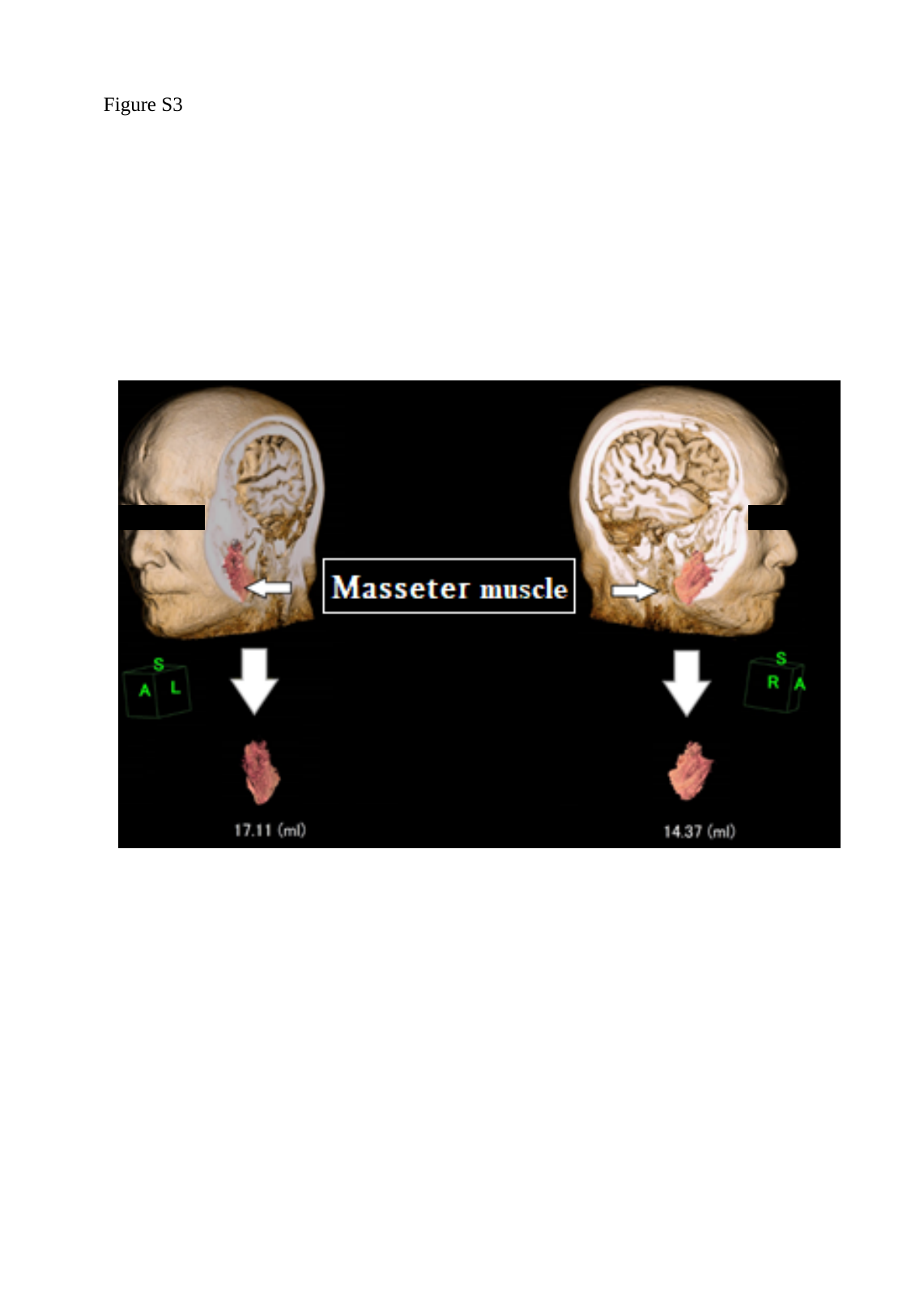

Figure S3
